# Development, Validation, and Implementation of a Stress Management Intervention for Rescue Workers in Rawalpindi: A protocol for a mixed-method study

**DOI:** 10.64898/2026.05.09.26352786

**Authors:** Iram Yasir, Ikhlaq Ahmed, Umar Farooq Bhatti, Shahzad Ali Khan, Abid Malik

**Author notes:** Corresponding Author: *Iram Yasir.

## Abstract

**Introduction:** Occupational stress among rescue workers is a major global public health concern. Rescue workers, including paramedics, firefighters, and disaster response teams, are consistently exposed to traumatic events, long working hours, physical hazards, and emotionally charged situations. These chronic stressors make them one of the most vulnerable groups to psychological distress, burnout, anxiety, depression, and post-traumatic stress disorders. In the local context of Pakistan, workplace mental health remains a neglected area. Despite stress and burnout being widely reported in these sectors, limited evidence-based interventions are available. Therefore, the study aims to develop and evaluate a locally tailored intervention to improve the mental health and psychosocial well-being of rescue workers.

**Objectives:**

1. To develop a culturally appropriate stress management intervention to promote mental health for rescue workers in Rawalpindi.
2. To validate the content and structure of stress management intervention for rescue workers
3. To evaluate the effectiveness of stress management intervention for rescue workers in Rawalpindi

**Method:** The ethical approval of the study has already been obtained from the ethical review board of Health Services Academy (00013/HSA/PhD-2022) and Rescue 1122 District Headquarters, Rawalpindi. Data will be collected after obtaining informed written consent from relevant stakeholders. Data collection will start from April 2026 and will be completed in six months. Data compilation and results are expected by December 2026. Data collection will involve a scoping review to explore stress determinants and intervention components, and then a qualitative phase in which data will be collected through focus group discussions from potential Stakeholders (rescue workers, mental health experts, and program managers) to identify and validate stress determinants. Triangulation of data will be done to integrate qualitative findings with findings from the review. In the second phase, validation will be done by intervention development experts. The third phase aims to evaluate the effectiveness of the developed intervention using a quasi-experimental pre-post design. A total of 154 participants evaluated with the Perceived Stress Scale Score will be employed through a stratified sampling technique. The primary outcome is defined as remission from stress at 3 months, measured with the PSS.

**Discussion:** It is anticipated that the study will result in the development of a culturally appropriate and evidence-based stress management intervention for rescue workers, thus contributing to sustainable improvement in rescuers’ mental health and job performance.

## Introduction

Globally, the frequency and intensity of disasters have increased enormously due to climate change, urbanization, and technological risks^1^. The increasing complexity and magnitude of disasters have necessitated the development of specialized emergency response systems and trained personnel capable of providing medical assistance and disaster management services. These frontline responders, i.e., paramedics, firefighters, emergency medical technicians, and rescue officers, are often the first to arrive at scenes of devastation, however exposed to severe human suffering, mass casualties, and loss of life^2^. Repeated exposure to traumatic events can make them one of the most vulnerable groups to a spectrum of psychological problems^3^.

This vulnerability is consistent with broader evidence indicating that individuals working in high-stress environments, such as healthcare providers, humanitarian workers, law enforcement personnel, and emergency responders, are consistently exposed to chronic stressors that can adversely affect their mental health^4^, decision-making^5^, and overall well-being^6^. Prolonged stress in such professions not only leads to burnout, anxiety, and depression^7^ but also compromises the quality of services provided to communities^8^.

In Pakistan, recurrent emergencies such as earthquakes, floods, frequent road traffic accidents, and terrorism-related incidents have tested the endurance of emergency response emergencies, particularly Rescue 1122, the country’s leading pre-hospital emergency service. A recent study from Khyber Pakhtunkhwa revealed elevated levels of stress, sleep disturbance, and emotional exhaustion among frontline emergency workers, highlighting the urgent need for structured psychological support systems^9^. Disaster-exposed responders also report higher prevalence of physical symptoms such as fatigue, headaches, and gastrointestinal problems^10^. Additionally, occupational stress has been associated with reduced cardiorespiratory fitness and poor sleep quality, which further increases the risk of chronic diseases among rescue workers^11^.

The growing concern regarding the mental well-being of emergency responders has also been recognized nationally. It calls for a multidisciplinary approach for an integration of mental health services in all emergency preparedness and response program^12^. Empirical evidence further supports these concerns. A study conducted by Martinez revealed that 9.8% PTSD prevalence among rescue workers, and it was found same compared with earlier findings from 10 years ago^13^. Similarly, research conducted in Rahim Yar Khan showed that 32.6% and 45.7% of rescue workers of 1122 are experiencing severe to extremely severe levels of depressive and anxiety symptoms^14^. Another study conducted on 1122 rescue workers in Lahore found more than 33% of the participants to be suffering from psychological distress^15^.

Occupational stress among rescue workers is a significant global public health challenge. The World Health Organization (WHO-2020) and International Labor Organization (ILO-2022) have recognized occupational stress as a growing threat to mental health and productivity^16^,^17^. Unmanaged stress not only affects individual well-being but also has a direct impact on the quality of emergency response, decision-making, and public safety outcomes^18^. Evidence from high-income countries indicates that elevated stress among emergency responders is associated with absenteeism, high turnover, substance misuse, and diminished operational efficiency^19^. The World Health Organization (2025) recognizes mental health and psychological support MHPSS as a core component of effective emergency response, emphasizing that the psychological well-being of rescuers is integral to overall disaster resilience and recovery^20^.

While numerous stress management interventions such as cognitive behavioral therapy, resilience training, and mindfulness-based stress reduction have demonstrated efficacy in improving psychological outcomes among first responders in high-income settings, their applicability in low and middle-income countries remains limited^21,22^.

In the local context of Pakistan, workplace mental health remains a neglected area. Individuals working in a stressful environment have limited access to a context-appropriate psychological support system^23,24.^ Routine mental health care is often inaccessible due to stigma, time constraints, and limited services^25^. Despite stress and burnout being widely reported in sectors such as healthcare, education, and corporate environments, however, limited evidence-based interventions are available^26,27^. Evidence shows that there is significance in introducing culturally relevant non-specialized interventions, and they should be incorporated into workplace settings^28^.

Developing a locally tailored intervention has high cultural fit as it addresses unique stressors in a targeted and specific way. It may also ensure deeper effectiveness across mental health, performance, and well-being domains. It will not only bridge the gap but will also generate insights for broader adoption and implementation at scale^29^. Such an intervention will improve individual health outcomes but also contribute to the efficiency and sustainability of vital public services^28^. By integrating elements of emotional intelligence, stress reduction strategies, and workplace support, this intervention will have the potential to empower professionals, reduce mental health stigma, and foster healthier work environments in resource-constrained settings like Pakistan, thus contributing to the global discourse on occupational mental health. Thus, keeping in mind the above-mentioned status, this mixed-method study is designed to develop and evaluate a Stress Management Intervention for Rescue Workers in Rawalpindi.

## Aim

To improve the mental health and psychosocial well-being of rescue workers by developing and evaluating the effectiveness of Stress management interventions.

## Objectives

1. To develop a culturally appropriate stress management intervention to promote mental health for rescue workers in Rawalpindi.
2. To validate the content and structure of stress management interventions for rescue workers.
3. To evaluate the effectiveness of stress management intervention for rescue workers in Rawalpindi

## Method

### Conceptual framework

The present study will use the Medical Research Council (MRC) framework, and selected components will be used according to the scope of the study and availability of resources. The MRC framework is ideal for a mixed-method study where qualitative data inform design and quantitative data test outcomes. It provides a rigorous, stepwise, and theory-informed approach specifically tailored for complex health interventions.

The Medical Research Council (MRC) Framework (Fig 1) for the development of complex interventions has been widely applied across disciplines, which suggests its flexible, non-linear approach may be usefully applied to the iterative design processes used in the development of new systems^33^. However, capturing all the interacting components and outcomes of complex interventions can be methodologically challenging.

**Fig 1:**
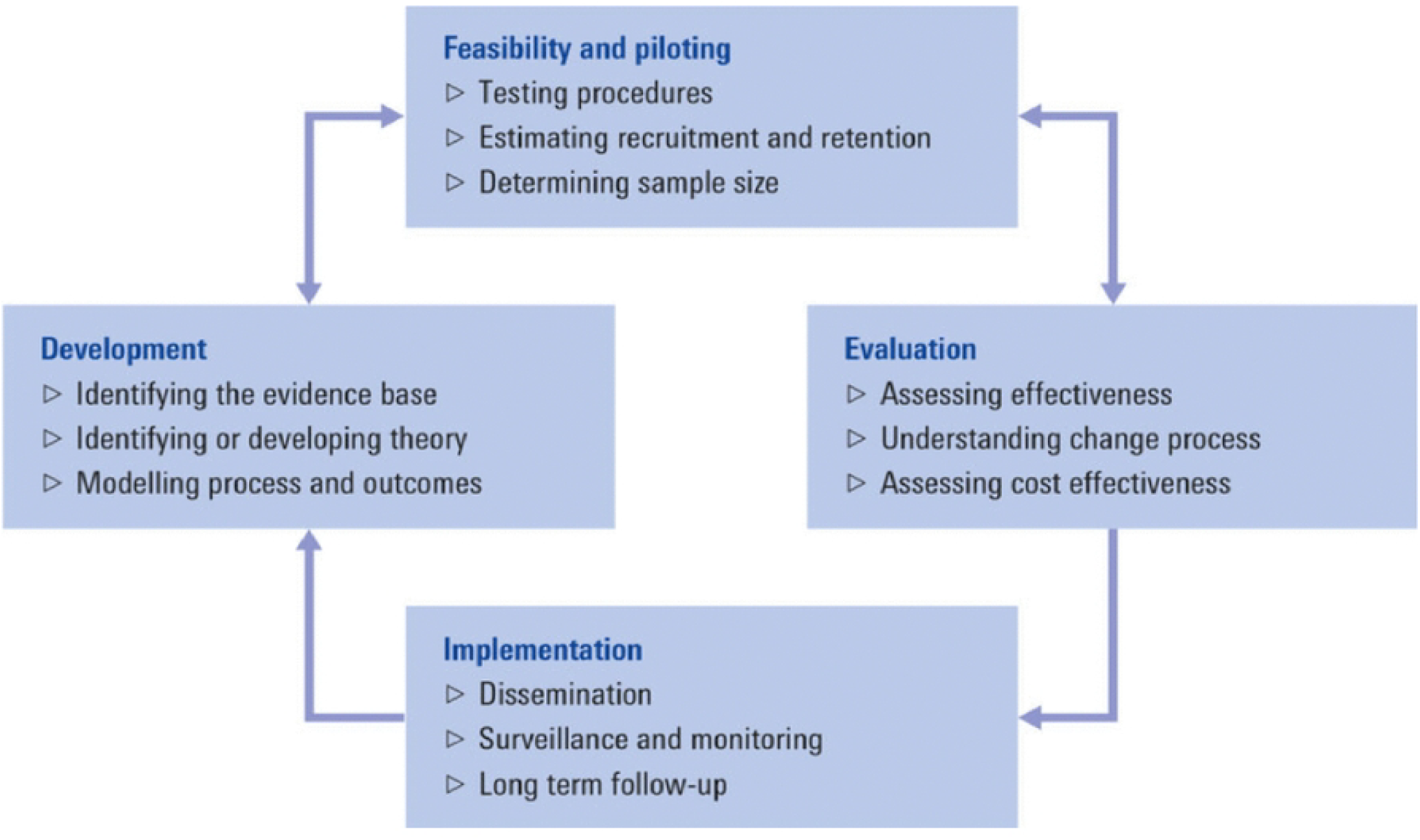
The Medical Research Council (MRC) Framework

We will be using the SPIRIT checklist annotated with TRENDS while writing our protocol. The proposed study comprises three phases. Methods for each work phase are presented below:

### PHASE 1: DEVELOPMENT

#### Scoping review

The primary goal of this phase is to map the existing evidence on stress determinants and stress management interventions applicable to humanitarian work environments. This will be done by conducting a scoping review. We will include PubMed, Taylor and Francis, Cochrane Library databases, and Google Scholar

##### Data Analysis Plan

Determinants of stress and effectiveness data for management interventions will be extracted. Extracted information will not only inform the development of the topic guide for the conduction of Focus group discussions, but also the intervention components will be identified.

#### Qualitative Data

This will be collected by conducting a Focus Group Discussion with various stakeholders.

##### Study population

It will include rescue workers (Ambulance drivers, Paramedics, and Fire fighters), Program Managers of 1122, and Mental Health experts (Psychiatrists and Psychologists)

Three different topic guides for various stakeholder groups will be developed to identify context-specific stressors, organizational context, and mental health needs that will guide the development of the intervention.

##### Data collection

The Focus group discussion will be conducted either in English or Urdu, as per the participants’ preference. Face-to-face or virtual format will be carried out. Face-to-face sessions at a location chosen by participants and virtual sessions via Zoom or Microsoft Teams. Duration will range from 30 to 60 minutes. The focus group discussion will be conducted by two trained research assistants. Recruitment has not yet started. Data collection will start from June 2026 and will be completed in nine months. Results expected by June 2027

##### Data analysis plan

Transcripts will be read several times in order to fully comprehend the perspectives of the participants. Data analysis will be conducted using reflective thematic analysis. This will validate determinants and coping strategies.

#### Develop an Intervention to address Stress

Findings from the scoping review, as well as Focus Group Discussions, will be utilized to identify and design the initial draft material for intervention regarding content, format, duration, and intensity as follows:

1. Outline of the modules or sessions and core components of the intervention. The intervention will likely comprise the following psychological techniques based on scoping review and qualitative data collected:
  - Understanding stress and occupational triggers;
  - Stress management skills such as diaphragmatic breathing, progressive muscle relaxation, and grounding;
  - Cognitive restructuring to challenge and reframe negative thoughts;
  - Peer support and communication strategies;
  - Self-care and resilience-building practices; and
  - Learning problem management
2. Development of potential delivery methods (e.g., individual sessions, group workshops, online modules, blended approaches).
3. Duration and intensity of the intervention.
4. Principles/guidelines for tailoring the intervention to specific workplace contexts.

### PHASE 2: VALIDATION

The goal of this phase is to establish content and face validity with experts and users.

#### Face validity and content validity (Expert Consultation)

It will be ensured by engaging 4-8 experts in stress management, occupational health, and intervention development to review the proposed framework and intervention materials. Feedback will be gathered on the relevance, feasibility, and potential effectiveness of the developed intervention by conducting in-depth interviews. Expert feedback will be incorporated to refine the intervention framework and intervention materials.

#### Pilot Testing of Intervention Materials

We will conduct a small-scale pilot test of the draft intervention materials with a small sample (15-20) of individuals at the Gujar Khan rescue 1122 center to assess feasibility, acceptability, and initial impact. 3-6 cognitive interviews from participants regarding acceptability, comprehension, and cultural fit will be utilized to finalize the intervention.

#### Finalize Intervention Protocol and Materials

Based on expert feedback and pilot testing, an intervention protocol and all associated materials will be finalized. This will serve as the basis for the implementation and evaluation in Work Package 3.

### PHASE 3: IMPLEMENTATION AND EVALUATION

This work package will include

#### Train the Trainers (TOTS) certificate program

The trainers will be trained by qualified mental health professionals, including clinical psychologists and researchers involved in intervention development, through a structured Training of Trainers (TOTS) program. A tool, ENACT, will be used for the selection of trainers to ensure high-quality, consistent, and effective delivery of an intervention. This tool measures a set of therapeutic competencies required for the effective psychological intervention, including delivery by non-specialists^31^.

First, eligibility criteria will be verified through a curriculum vitae review and verification of professional experience. Second, suitability will be assessed using the tool ENACT. Only trainers meeting cutoff thresholds proceeded to the final step, which involved attending a two-day standardized training workshop on intervention delivery, cultural sensitivity, and fidelity monitoring procedures.

#### Implement the intervention and conduct a quasi-study for preliminary evaluation

##### Selection of the Participants

###### Inclusion criteria

1. Individual with a high score on Perceived Stress Scale (PSS>14).
2. Those are intended to stay in the same organization for the study period.

###### Exclusion criteria

1. Individual with serious physical health complications (e.g Diabetes mellitus complications, Metabolic complications).
2. Individuals already diagnosed with any Psychiatric/psychological condition and receiving treatment for common mental disorders.

##### Implementation Site

Rescue 1122 is established under the PES Act 2006 for professional management of emergencies. It is working in 37 out of 41 total districts of Punjab. Organizations. Rescue 1122 Rawalpindi has a staff of 650 personnel, including 567 operational and 83 administrative staff.

As the headquarters of emergency response, situated at Murree Road, Rescue 1122 Rawalpindi District HQ faces frequent exposure to emergencies such as road traffic accidents, fires, natural disasters, and terrorist attacks. These high-risk and high-pressure scenarios make rescue workers in the city particularly vulnerable to stress, thus making it a suitable site for the implementation of research.

##### Study Design

A quasi-experimental study.

##### Sample Size

- Using the G Power sample size calculator and keeping the margin of error at 5% and power of 0.80, the sample size came out to be 128.
- Keeping in mind attrition loss, non-response, and loss to follow-up (20%), the final sample size is calculated to be around 154.

##### Sampling

We will employ proportionate sampling as follows:

- Ambulance drivers-202/567(35%) …54
- Paramedics-212/567(37%) 57
- Fire fighters-154/567(27%) 43

##### Data Collection and Outcome Measures

The data collection tool and study outcomes have been detailed in Table 1

**Table 1:** Schedule of study assessment activities for main study.

| Outcome | Measured/<br>assessed by | 3-month post-intervention |  | Translation and<br>Validity | Description of<br>outcome<br>measure |
| --- | --- | --- | --- | --- | --- |
|  |  | Primary<br>outcome | Secondary<br>outcome |  |  |
| Stress | PSS-10 | % of stress<br>reduction | x | Validated tool<br>used in stress<br>intervention<br>research | Self-reported:<br>10 items; 5-point scale;<br>categories of<br>severity |
| Anxiety | GAD-7 | x | Total mean<br>score | Screen for and<br>measure<br>generalized<br>anxiety disorders | DSM-IV<br>criteria-based<br>anxiety<br>screening tool:<br>7 items: 0-21<br>score;<br>categories of<br>severity |
| Depression | PHQ-9 | x | Total mean<br>score | Screen for and<br>measure severity<br>of depression | DSM-IV<br>criteria-based<br>depression<br>screening tool:<br>9 items: 0-27<br>score;<br>categories of<br>severity |

**Table 2:** Schedule of interventions development, enrolment, and assessment.

|  |  | STUDY PERIOD |  |  |
| --- | --- | --- | --- | --- |
| Timepoint |  | Baseline | Enrolment<br>t1 | 3 months<br>post-intervention |
| INTERVENTIONS |  |  |  |  |
| Developed intervention |  | x |  |  |
| ENROLMENT: |  |  |  |  |
| Eligibility screen |  |  | x |  |
| Informed consent |  |  | x |  |
| Baseline assessment |  |  | x |  |
| ASSESSMENTS |  |  |  |  |
| Diagnosis |  |  | x |  |
| Outcome assessment |  |  |  | x |

###### Baseline

Those who screen positive on PSS will be invited to participate in baseline and enrollment. A detailed questionnaire will be administered to all participants. This includes the demographic section, PSS, PHQ-9, and GAD-7.

###### Post-intervention follow-up

We will administer the same tools used at baseline and at post-intervention follow-up after 3 months.

###### Primary outcome

Our Primary outcome is defined as improvement in PSS score at 3 months after intervention. Perceived Stress Scale -PSS is a widely used psychological tool to measure how stressed a person feels over the last month in stress management research

###### Secondary Outcome

We will also collect data to screen anxiety and depression using the validated questionnaire GAD-7 and PHQ-9.

#### Data Analysis

Data will be analyzed by using SPSS version 28. Regarding socio-demographic variables, quantitative variables will be analyzed by calculating mean and standard deviation, while qualitative variables will be analyzed using frequencies and percentages. Paired T-test will be used to compare the mean scores before and after the intervention. Mediation Analysis will be conducted to examine whether the effect of the stress management intervention on psychological outcomes (e.g., stress, anxiety) was mediated by changes in variables such as coping strategies, self-efficacy, or perceived social support.

#### Data monitoring

The principal investigator will be responsible for reviewing and assessing recruitment, monitoring of safety and effectiveness, conduct, and data management.

## Data Availability

For Study Protocols: No datasets were generated or analysed during the current study. All relevant data from this study will be made available upon study completion

## Integration of Data

Integration will be done at various stages of study. Findings from the scoping review and qualitative interviews will be triangulated within phase I. The findings will directly feed into the design of the stress management intervention. Finally, after phase III, all the insights into how findings from each stage align to explain stress determinants, intervention design, and impact will be combined. A sample joint display is illustrated as a procedural diagram. Fig 2

**Fig 2:**
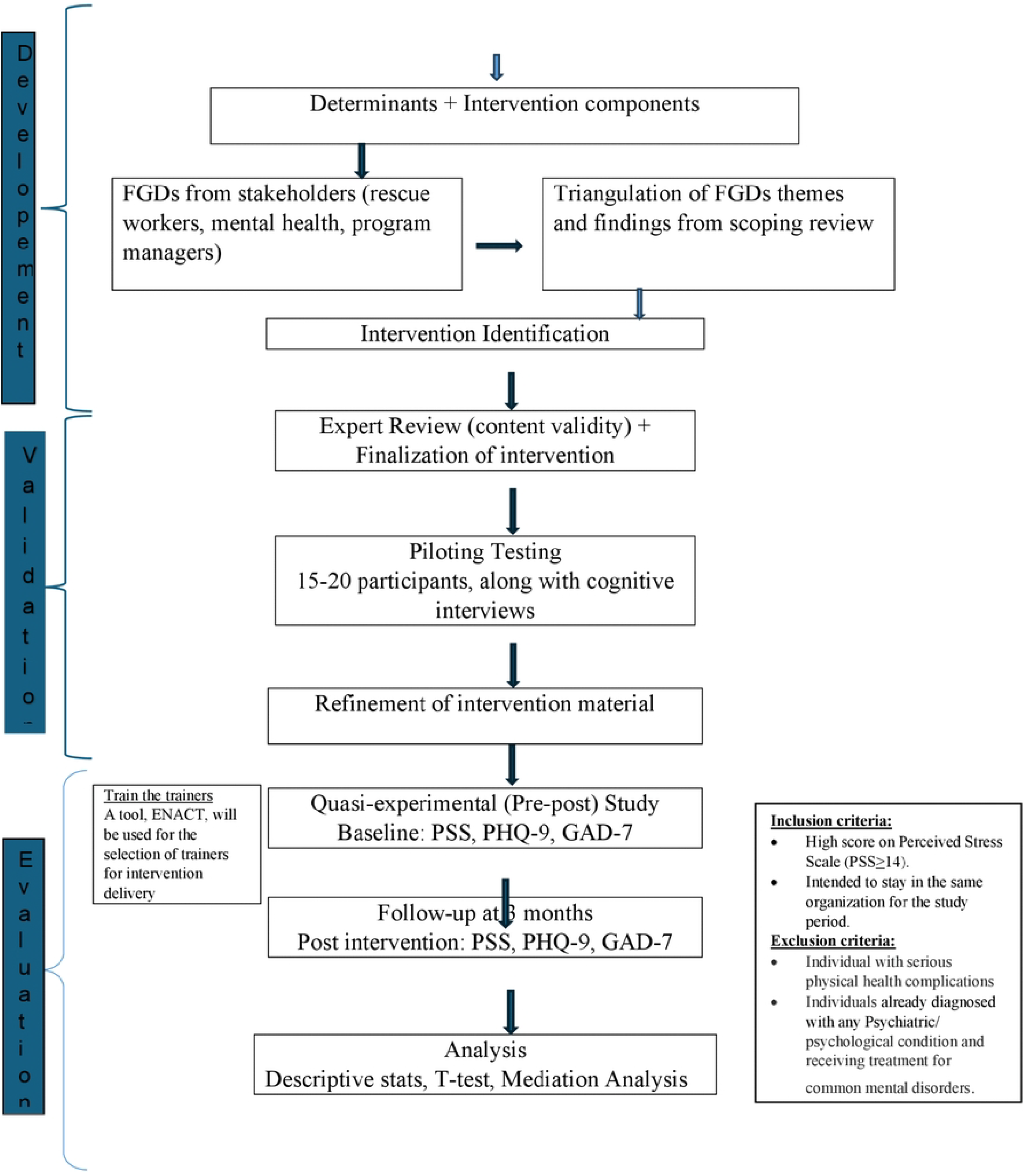
STUDYFLOW CHART

## Ethical considerations

Ethical clearance has been obtained from the Institutional Review Board of Health Services Academy (00013/HSA/PhD-2022) as well as from Rescue 1122 District Headquarters, Rawalpindi. Voluntary recruitment and written informed consent will be taken after informing about the protocol of the study. A copy of the information sheets and consent forms will be left with the participants. Informed consent will be sought from all participants before screening, baseline, and intervention delivery. Privacy of participants will be ensured. Data will be anonymous and confidential. Data will be kept under lock and key, and the principal investigator will be responsible for the security of the data. Participants will have the right to withdraw at any time in time. Adherence to the ethical guidelines of the relevant professional association and regulatory bodies will be ensured.

## Expected outcome

It is anticipated that the study will result in the development of a culturally appropriate and evidence-based stress management intervention for rescue workers. This intervention will be feasible for integration into existing rescue service training and support systems, contributing to sustainable improvement in responder mental health and job performance.

## Risk assessment and mitigation

A register will be maintained of potential adverse events, defined as any unfavorable signs or symptoms. For this study, an adverse event could be a panic attack, sleep problem, chest pain, body aches, or hospital admission of a participant due to any physical or psychiatric problem. Participants screened with anxiety and depression during the course of the study will be referred to the Institute of Psychiatry, Rawalpindi Medical University. It is a public healthcare facility providing specialized mental health services. This referral pathway will serve as a part of routine care support for participants. Those participants will be discontinued from the study altogether.

## Dissemination

Policy brief: Findings could advocate for health policies by highlighting the need for integration of stress management interventions for frontliners working in high-intensity situations.

Journal and Conference presentation: We will disseminate the findings through publications in open-access peer-reviewed journals having a large readership, as well as presentations at local, national, and international conferences

Stakeholder workshop: to promote opportunities for collaboration with stakeholders to promote the mental health and well-being of rescue workers

Social media: Social media campaigns on Instagram and Facebook from the webpages of the Public Health at Health Services Academy and Isra University.

## Supplementary Information

## Acknowledgement

Not Applicable

## Authors’ contributions

**Conceptualization:** Iram Yasir

**Methodology:** Iram Yasir

**Project administration:** Ikhlaq Ahmad

**Software:** Umar Farooq Bhatti

**Supervision:** Abid Malik

**Writing – Original draft:** Iram Yasir

**Writing-review & editing:** Shahzad Ali Khan

